# Correlation between autistic traits and brain functional connectivity in preschoolers with autism spectrum disorder: a resting state MEG study

**DOI:** 10.1101/2024.01.26.24301652

**Authors:** Matilde Taddei, Pablo Cuesta, Silvia Annunziata, Sara Bulgheroni, Silvia Esposito, Elisa Visani, Alice Granvillano, Sara Dotta, Davide Sebastiano Rossi, Ferruccio Panzica, Silvana Franceschetti, Giulia Varotto, Daria Riva

## Abstract

**Background:** Neurophysiological studies recognized that Autism Spectrum Disorder (ASD) is associated with altered patterns of over- and under-connectivity. However, few results are available about network organization in children with ASD in the early phases of development and the correlation with the severity of core autistic features.

**Methods:** The present study aimed at investigating the association between brain connectivity derived by MEG signals and severity of ASD traits measured with different diagnostic clinical scales, in a sample of 16 children with ASD aged 2 to 6 years.

**Results:** A significant correlation emerged between connectivity strength in cortical brain areas implicated in several resting state networks and the severity of communication anomalies, social interaction problems, social affect problems, and repetitive behaviors. Seed analysis revealed that this pattern of correlation was mainly caused by global rather than local effects.

**Conclusions:** The present results confirm that altered connectivity strength in several resting state networks is related to clinical features and may contribute to neurofunctional correlates of ASD. Future studies implementing the same method on a wider and stratified sample may further support functional connectivity as a possible biomarker of the condition.

## 1. Introduction

Autism spectrum disorder (ASD) is a neurodevelopmental condition characterized by early onset problems in social behaviours and communication, restricted interests, and repetitive patterns of behaviours (American Psychiatric Association, 2022).The disorder typically reveals in the first years of life with a wide range of clinical presentations and persists throughout life. The etiological factors of ASD remain largely unknown, but many studies are approaching the etiology of the disorder from different points of view, looking at immunity, epigenetics, environmental factors, and genetics (Masini et al., 2020) and investigating several behavioural, neuroanatomical, and functional endophenotypes [1] The diagnostic work-up resides on the evaluation of the child’s behavioural characteristics and is based on scored parent’s reports of the present and past behaviour, supported by structured play sessions designed to elicit current behavioural features [2].

Many studies investigated the presence of functional differences in brain networks suitable to understand the pathophysiology of ASD and to identify possible diagnostic and prognostic biological markers. Different functional measures based on neurophysiological tests or brain imaging have been applied to evaluate either the response to specific stimuli or the resting state signals [3]. Using whole-brain electroencephalography (EEG) and magnetoencephalography (MEG) recordings, previous studies recognized that ASD is associated with altered patterns of over- and under-connectivity [4]. Various observations supported long-range under-connectivity in ASD, namely in low-frequency bands, while the characteristics of local connectivity remain unclear, with a tendency for overconnectivity in high-frequency ranges [5], [6].

Various EEG studies have been performed in infants and toddlers ([7] for a review) with the aim of detecting predictive electrophysiological markers in subjects with high likelihood of being diagnosed with autism. When specifically focusing on brain connectivity, most EEG/MEG studies included teenagers or older children [5], [8]–[10], therefore possibly capturing the outcome of protracted cortical or subcortical reorganization processes, while only a few studies were performed in early childhood [11], the period of life in which the full-blown manifestations of the condition overtly appear. In general, studies investigated children with ASD without developmental delay or intellectual disability [5], [9], [12] to facilitate comparison with typically developing children, but leading to results less representative of the whole ASD spectrum, which involves at least 37.9% of children with intellectual impairment [13]. Some case-control studies found also that an altered resting state neurophysiological connectivity in high-frequencies ranges, in particular underconnectivity and reduced network efficiency in beta and gamma bands, is related to higher levels of autistic traits reported by parents [9], [12]. However, these studies involved school-age children and provided an association with traits severity assessed by screening questionnaires, while the association with measures derived by diagnostic instruments, such as Autism Diagnostic Observation Schedule-2nd Edition (ADOS-2) or Autism Diagnostic Interview-Revised (ADI-R), is less investigated and provided poor results [12].

Compared to screening questionnaires, the direct observation of the child by a specifically trained clinician through ADOS-2, integrated with the information obtained by ADI-R clinical interview, has provided the highest diagnostic stability and reliability [14] and is advisable, especially in young children at first diagnostic assessment.

With the aim of facing the neurophysiological network characteristic in children with ASD in the early phases of development, we performed the present study in a population of children (aged 2 to 6 years) unaffected by brain damage or developmental encephalopathies and still not exposed to psychotropic medications.

The main objective of the study was to identify if resting state MEG network-based parameters correlate with the severity of autistic traits, as measured with different diagnostic clinical scales, and could be thus considered as possible future biomarkers for ASD early diagnosis.

## 2. Material and methods

### 2.1 Participants

We selected a group of preschoolers with a diagnosis of ASD admitted to the Unit for Neurogenetic syndromes with Intellectual Disabilities and Autism Spectrum Disorders of the IRCCS Carlo Besta. Exclusion criteria were: the presence of major dysmorphic features and malformations, the presence of brain structural alterations by Magnetic Resonance Imaging (MRI), and diagnosis of epilepsy or epileptic syndromes. ASD diagnosis was made in accordance with DSM 5 criteria and confirmed by ADI-R [15] and ADOS-2 [16].

Further information about sample selection is provided in supplementary materials. The final sample is formed by 16 children (median age 39.25 months, range 30-77 months), 3 females and 13 males. All the examinations were performed with the written informed consent of the subject’s parents. The study was performed according to the Declaration of Helsinki and was approved by the Ethics Committee of Fondazione IRCCS Istituto Neurologico Carlo Besta.

### 2.2 Clinical scales and exams

ADI-R is a clinical tool for the diagnosis of ASD in children and adults consisting of a standardized and semi-structured clinical interview for caregivers. New algorithms have been developed for toddlers and young preschoolers from 12 to 47 months of age [17]. For the aim of the present study, we calculated three severity indices, one for each functional domain investigated by ADI-R: *ADI Interaction*, describing the quality of reciprocal social interaction, *ADI Communication* about child’s language and non-verbal communication anomalies, and *ADI Interests* describing the presence of restricted, repetitive, or stereotyped interests or behaviors.

ADOS-2 is a semi-structured evaluation of play used with children and adults suspected for ASD. For the aim of the present study, we have calculated two separated indices, respectively for the social affect (ADOS Social Affect) and restricted and repetitive behaviors (ADOS Restricted and Repetitive Behaviours) domains. The mean for each ADOS-2 and ADI-R domain was obtained by dividing the total score by the number of items included, to accommodate for differences in the modules and diagnostic algorithms, which are differentiated depending on developmental and language levels. Higher scores reflect higher severity of traits in terms of pervasiveness and intensity. Information about clinical and instrumental evaluation of the sample are provided in supplementary materials.

### 2.3 MEG Data acquisition

All patients underwent MEG in awake condition, after administration of light sedation (Melatonin + Hydroxyzine or low dose of Chlorpromazine). The acquisition took place in a supine lying position with the presence of the caregiver of the children inside the magnetic shielded room. Patients were instructed to relax, remain calm, and close their eyes. The MEG signals were acquired using a 306-channel whole head MEG system (Triux, MEGIN, Helsinki, Finland), at 1Ks of sample frequency. Bipolar electro-oculographic (EOG) and electrocardiographic signals (ECG) were also acquired. The participant’s head position inside the MEG helmet was continuously monitored by five head position identification (HPI) coils located on the scalp. The locations of these coils, together with three anatomical landmarks (nasion, right and left preauricular), and additional scalp points were digitized before the recording by means of a 3D digitizer (FASTRAK, Polhemus, Colchester, VT, USA). At least 15 minutes of MEG signal was recorded for each child.

To allow MEG-MRI co-registration, for each patient an isotropic 1mm3 volumetric T1-weighted sequence was also acquired.

### 2.4 MEG Data analysis

#### 2.4.a Signal pre-preprocessing

The detailed pre-processing procedure and the source reconstruction have been reported in detail in [18]. Temporal signal space separation (tSSS) [19] was applied to reduce the noise in the MEG data using MaxFilter 2.2 software (Megin Oy). Magnetometers data were automatically scanned for ocular, muscle, and jump artefacts using the Fieldtrip software [20], which were visually confirmed by an MEG expert (SF). The remaining artefact-free data were segmented in 4 seconds segments (epochs), plus 2 seconds of real data at each side as padding. Finally, MEG time series were filtered into delta (2-4Hz), theta (4-8Hz), alpha (8-12Hz), beta (12-30 Hz) and gamma (30-45 Hz).

#### 2.4.b Source Reconstruction

We used a regular grid of 1 cm spacing in the Montreal Neurological Institute (MNI) template and labelled according to the Automated Anatomical Libelling (AAL) atlas [21]. This source model consisted of 1202 positions in 78 cortical areas and was transformed to subject space using a linear transformation between the template and the T1-weighted MRI of the participant. The forward model solution used a single-shell method [22] with a unique boundary defined by the inner skull extracted from each individual T1 image. Source reconstruction was carried out independently for each subject and frequency band with a linearly constrained minimum variance (LCMV) beamformer [23]. This method has yielded reliable results for the estimation of resting-state functional connectivity (FC) [24]. FC between all 1202 nodes was assessed, for each participant and frequency band, by computing the phase locking value (PLV) [25], This metric was chosen for its proven high reliability across sessions, a key property of connectivity measures, guarantying repeatability and consistency of single-subject and group level results [26]. Lastly, we computed the nodal strength (also known as weighted global connectivity), which is defined as the sum of its FC with the rest of the nodes. To account for the number of links, the strength of each node was then normalized by dividing the number of links connected to it. This procedure resulted in one brain map of normalized node strengths (henceforth named FC-st) per each participant and frequency band.

### 2.5 Statistical Analyses

#### 2.5.a Functional Connectivity Strength (FC-st) analyses

We tested the presence of significant partial correlation between FC-st values and behavioral scales (BS: ADOS-2 and ADI-R clinical scales). The assessment was based on a cluster-based permutation test (CBPT) described previously [27] and was applied independently for each frequency band. The units of study were sets (clusters) of spatially adjacent nodes that presented a significant partial correlation (Spearman correlation using age at MEG recording as a covariate, p< 0.05) in the same direction between FC-st values and each BS variable. In this framework, a cluster can be considered as a functional unit. Candidate clusters were required to have at least 1% of the total nodes (i.e. 12 nodes). Spearman rho values were Fisher Z-transformed. Cluster mass statistics were assessed through the sum of all z-values of the nodes belonging to the cluster. Then, to control for multiple comparisons, this procedure was repeated 5000 times after randomly shuffling the correspondence between FC-st and each BS measure across all participants. At each repetition, the maximum statistic of the surrogate clusters was stored, creating a maximal null distribution that ensured the control of the family-wise error rate (FWER) at the cluster level. The resulting cluster-based statistic (CBPT) *p*-value for a candidate cluster corresponded with the proportion of the permutation distribution with cluster-statistic values greater than or equal to the cluster-statistic value of the original data. Only those clusters that resulted significant (p < 0.05) after this step were considered in further analyses. Then, we used the average of the FC-st values of the members of the cluster to obtain a representative FC marker.

#### 2.5.b Seed-based analyses

To examine whether the FC-st results were caused by global or region-specific effects, we performed complementary seed analyses, using the previous clusters as seeds. In this seed-based correlation analysis, we identified the specific connections of each of the main clusters with the rest of the brain that were significantly modulated by severity scores at ADI-R and ADOS-2 scales. Specifically, the seed region was constituted by grouping all nodes located within a radius of 20 mm from the mass centre of the cluster. Then, the FC between each source position and the seed was computed by averaging the FC of the corresponding source position with all the nodes contained in the seed region. Using these new node-to-seed FC, additional cluster-based analyses were performed with the same parameters as the original analysis. Only clusters that did not overlap with the original seed-cluster were reported in this study. All statistical analyses were carried out using Matlab R2020b (Mathworks Inc).

## 3. Results

### 3.2 MEG Functional connectivity and association to Behavioural Scales

Overall, all the obtained results indicate the presence of a significant correlation between FC indexes and some scores of the ADOS-2 and ADI-R clinical scales. For all the obtained results, the significant clusters revealed a positive correlation, indicating that more severe behavioural problems are associated with higher FC-st. Tables S2 to S5 display the ROIs with significant correlations between FC-st and the different clinical scale. Only the Regions of Interest (ROIs) involving clusters with at least 30% of nodes displaying significant correlation are reported. Functional implications of each anatomical area (as defined by Automated Anatomical Labeling (AAL) atlas (Tzourio-Mazoyer et al., 2002) are also specified to ease the interpretation of the results, based on recent atlases of functional connectivity of well assessed resting state networks (Default Mode, Salience, Central Executive, Visual and Sensorimotor) (Doucet et al., 2019).

All the significant correlations involve beta and gamma bands, being the gamma band the most involved, while no relationship between clinical scales and lower frequency bands could be detected. Moreover, no correlation was found with the general developmental quotient.

#### 3.2.a FC and ADI-R scores correlation

Significant correlations were found between FC-st and all the three ADI-R subscales (Communication, Interaction and Interests).

ADI Communication scale correlates with FC-st in both beta and gamma bands. In the beta band, two clusters were identified: the bigger one (β_Com1) located in the right hemisphere and mainly involving parietal ROIs, and a second smaller cluster (β_Com2), located in the left superior parietal gyrus. Regarding the gamma band, a single and wider significant cluster (γ_Com) was detected, involving bilateral parietal, occipital and limbic ROIs. Results for ADI Communication are depicted in Figure 1 and Table S2.

**Figure 1.**
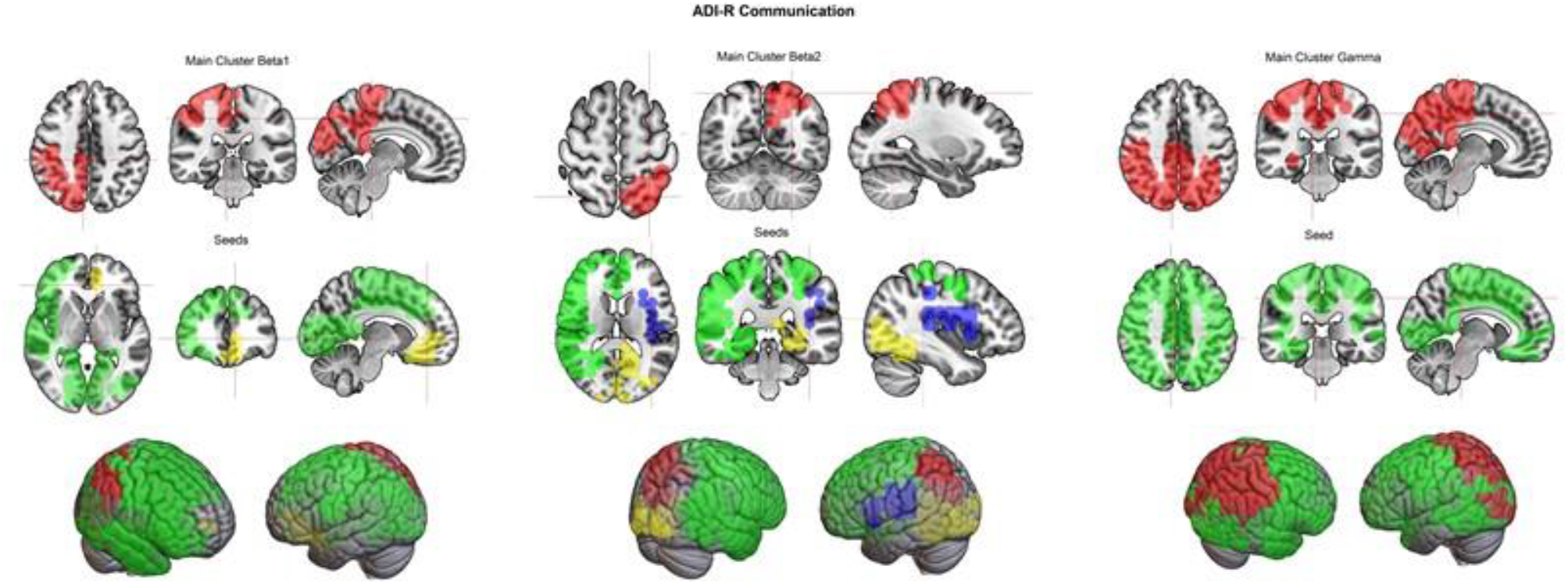
Main and Seed clusters for ADI Communication severity score. Figures 1 (upper side) shows the bidimensional representation of the FC-st main clusters for beta and gamma bands (in red color), superimposed on MNI anatomical space. Figures 1 (middle) represents seed clusters (in green, yellow and blue color) for each main cluster. Figures 1 (lower side) represents both the main and seed clusters for beta and gamma bands significantly associated with ADI Communication severity score superimposed on 3d whole brain. The right of the figure is the left of the brain.

ADI Interaction scores revealed a significant correlation with FC-st only in gamma band, where three different narrow clusters were identified: the first one (γ_Intera1) in right parietal ROIs, the second (γ_Intera2) in the left superior parietal gyrus and the right paracentral lobule, and a third (γ_Intera3) involving left occipital ROIs. (Figure 2 and Table S4).

**Figure 2.**
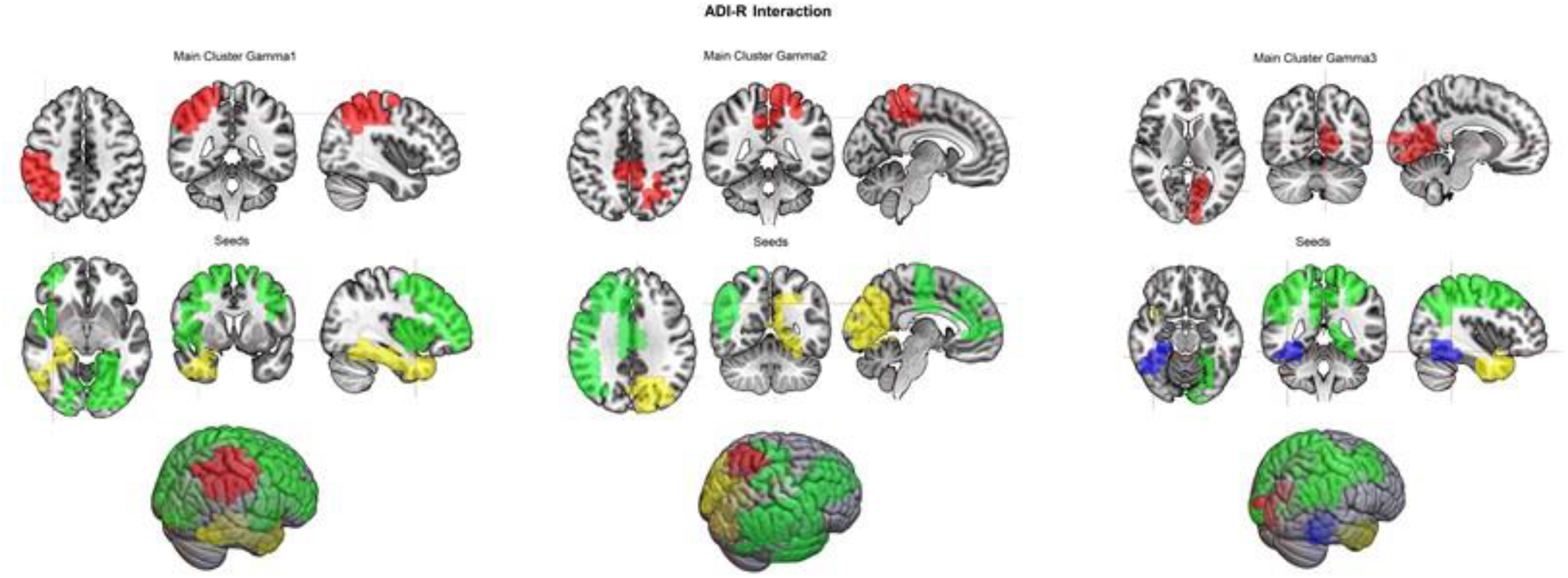
Main and Seed clusters for ADI Interaction severity score. Figures 2 (upper side) shows the bidimensional representation of the FC-st main clusters for gamma band (in red color), superimposed on MNI anatomical space. Figures 2 (middle) represents seed clusters (in green, yellow and blue color) for each main cluster. Figures 2 (lower side) represent both the main and seed clusters for gamma bands significantly associated with ADI Interaction severity score superimposed on 3d whole brain. The right of the figure is the left of the brain.

ADI Interests showed more localized and smaller clusters, two in beta and one gamma band, all limited to the left hemisphere. The beta band’s clusters involve left temporal and occipital ROIs (β_Ints1) as well as the left paracentral lobule (β_Ints2). Concerning gamma band, cluster of FC correlation only involved left temporal ROIs (γ_Ints) (Figure 3 and Table S4).

**Figure 3.**
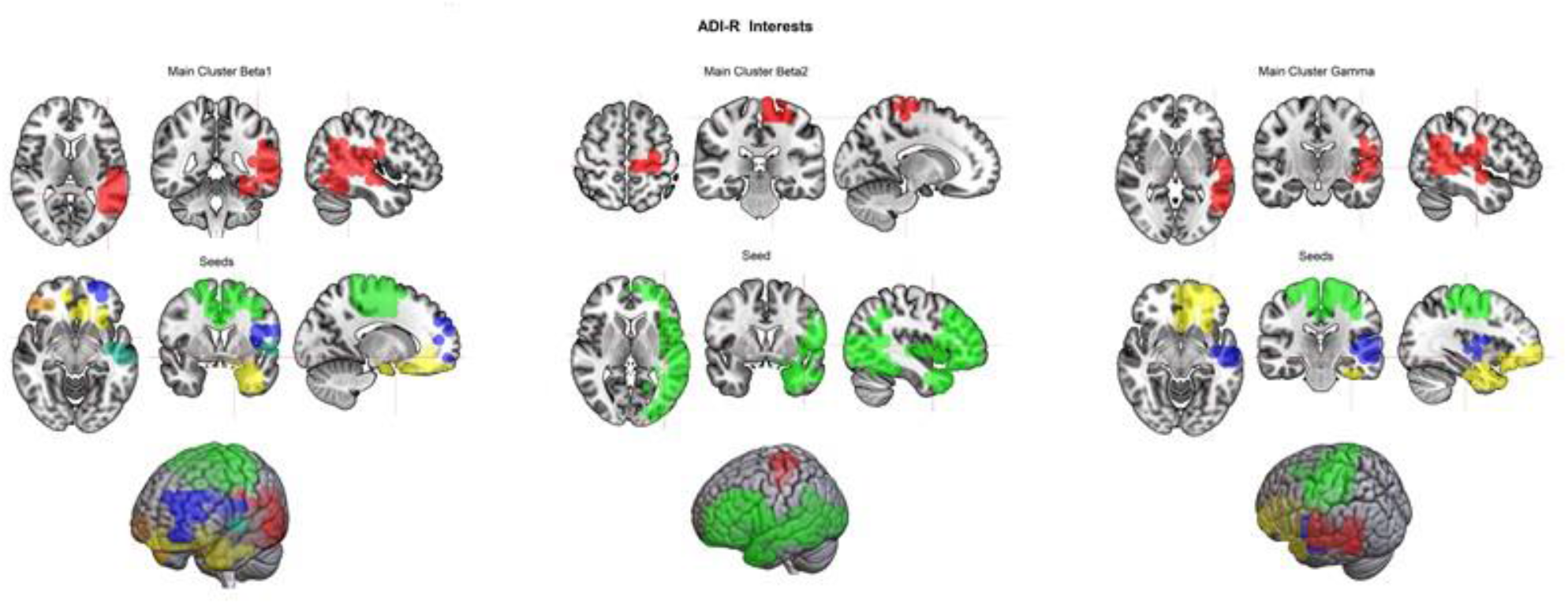
Main and Seed clusters for ADI Interests severity score. Figures 3 (upper side) shows the bidimensional representation of the FC-st main clusters for beta and gamma bands (in red color), superimposed on MNI anatomical space. Figures 3 (middle) represents seed clusters (in green, yellow, orange, light blue and dark blue color) for each main cluster. Figures 3 (lower side) represents both the main and seed clusters for beta and gamma bands significantly associated with ADI Interests severity score superimposed on 3d whole brain. The right of the figure is the left of the brain.

#### 3.2.b FC and ADOS-2 scores correlation

A significant correlation was found only with respect to the Social Affect subscale, with two localized, but bilateral, clusters in gamma band, one (γ_Soc1) involving right parietal ROIs (inferior parietal and supramarginal gyrus) and the second one (γ_Soc2) including left occipital ROIs, namely the lingual gyrus and the calcarine fissure (Figure 4 and Table S5).

**Figure 4.**
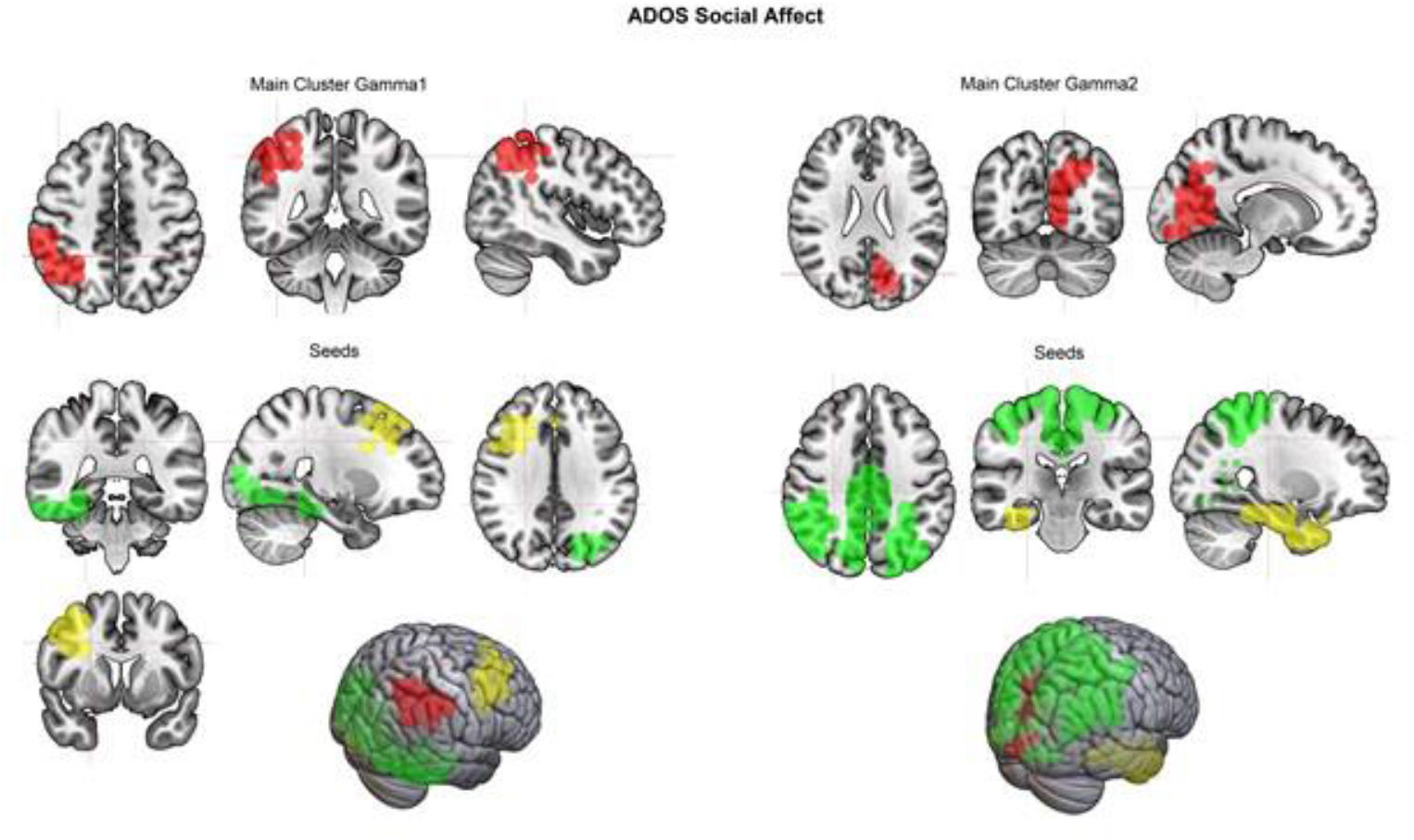
Main and Seed clusters for ADOS Social Affect severity score. Figures 4 (upper side) shows the bidimensional representation of the FC-st main clusters for gamma band (in red color), superimposed on MNI anatomical space. Figures 4 (middle) represents seed clusters (in green and yellow color). Figures 4 (lower side) represents both the main and seed clusters for beta and gamma bands significantly associated with ADOS Social Affect severity score superimposed on 3d whole brain. The right of the figure is the left of the brain.

For the others ADOS-2 subscales, i.e. Composite score and Restricted-repetitive behaviours, no significant correlation with FC-st alterations could be found, whereas the same trend of positive correlation between traits severity and FC alteration was present.

### 3.3 Seed results

Results of seed-based analyses for each main cluster and each behavioural domain are presented in Figure 1-4, Supplementary Tables S6-S9 and Supplementary Results. Briefly, seed analyses revealed that the increased connectivity was mainly related to spread connections. For ADI Communication, the involved ROIs were localized in several bilateral fronto-parietal, temporo-occipital and limbic ROIs, apart from two left-lateralized clusters for beta-bands in the frontal gyrus rectus, the inferior frontal gyrus and the occipital lobe (see Figure 1 and Supplementary Table S6). For ADI Interaction, many involved ROIs were located in left-lateralized or bilateral fronto-parietal, occipital and temporal regions, two clusters were right-lateralized cluster in the temporo-limbic regions and fusiform gyrus regions (see Figure 2 and Supplementary Table S7). For ADI Restricted Interests, many clusters were located in the fronto-parietal sensorimotor areas around the central sulcus but extend also to the left frontal, temporal and limbic regions; one cluster for beta band was located in the right inferior frontal gyrus (see Figure 3 and Supplementary Table S8). For ADOS Social Affect, the involved ROIs are mainly in the bilateral occipital, parietal and cingulate regions and in the right middle frontal gyrus, temporal lobe and parahippocampus (see Figure 4 and Supplementary Table S9).

## 4. Discussion

The present study was aimed at examining correlation between MEG functional connectivity and autistic traits to aid in early autism diagnosis. A significant correlation emerged between connectivity strength in cortical brain areas and severity of autistic traits derived by diagnostics ADI-R and ADOS scales. Higher levels of autistic traits positively correlated with higher gamma and beta bands connectivity in areas belonging to several resting state functional networks i.e. the Default mode, Central executive, Salience, Visual and Sensorimotor, according to recent consensus about the definition of the human connectome [28], [29]. Seed analysis revealed that the increased connectivity was mainly caused by global than by local effects. In fact, for all the clinical domains, functional coupling emerged between the posterior main clusters and several, not overlapping, second clusters, spread out in different brain regions encompassing frontal and limbic areas.

This evidence of a spread, mainly un-specialized overconnectivity related to ASD traits severity in an early phase of life and development, is in line with previous neuropathological and neurobiological data, that reported ASD as a condition resulting from overall brain altered reorganization beginning early in development [30]. Moreover, changes in brain intrinsic functional connectivity by fMRI was found to be related to autistic traits in individuals with ASD diagnosis from school-age throughout the lifespan [31] and also in population-based samples [32]. The correlation between an increased functional connectivity and traits severity, especially in young children, may be related to the reduced ability to differentiate functional networks and has been described as a compensatory mechanism to maladaptive differentiation of the cortical networks in people with ASD [6]. In this framework, the evidence of atypical connectivity in high frequency bands and in particular gamma band, received particular attention, primarily in evoked paradigms as considered to reflect sensory and attention processes, but also in spontaneous recordings, with evidence of altered connectivity when comparing ASD pre-schoolers to healthy controls [33]. Changes in beta-gamma connectivity have been reported also in other pathological conditions as a possible marker of (altered) brain functioning. For example, altered gamma connections have been observed in individuals with schizophrenia and auditory hallucinations [34], as well as atypical beta and gamma oscillations at rest have been reported in children and adults with Attention Deficit and Hyperactivity Disorder [35]. In fact, gamma oscillations and their network are thought to be fundamental for efficient cortico-cortical communication, and cognitive functioning [36]. In our study, the gamma (and to a less extent beta) functional connectivity essentially appears to positively correlate with the increased severity of autistic traits, in different social, communicative and behavioural functions explored by the different scales, in young children with ASD. Beta-gamma activity derives from a complex circuitry including inhibition and rebound excitation cycle mediated by the effects of GABAergic, fast-spiking interneurons acting on GABA-A receptors [37] and glutamatergic afferents at NMDA receptors, which provide excitatory input to these interneurons. Therefore, we can interpret the observed changes in this specific disorder and in a restricted age range as the enhanced activity within inhibitory circuitries connecting distant cortical areas. The present results confirm that an increased connectivity in several networks is related to higher social, communicative and behavioural problems and thus may contribute to neurofunctional correlates of ASD, as described also in multimodal fMRI-MEG study comparing children with autism and typically developing children [8], but also in fMRI studies investigating the correlation between intrinsic functional connectivity and autistic traits [31], [32]. It is worth noting that the present study revealed a significant correlation between severity measures and FC-st in occipital and posterior areas belonging to the visual, dorsal and ventral attention networks poorly described in previous reports on older children [9], [38]. Few studies described the correlation between autistic traits and MEG activity in gamma and alpha bands indices respectively in posterior temporal [10] and occipital [38] regions.

The lack of statistical correlation between severity measures and frontal regions’ connectivity does not exclude the involvement of the frontal area in our sample of ASD children, as also revealed by seed analysis results, which enhanced how frontal regions contribute to the main cluster’s increased connectivity. It could moreover suggest the role of a specific (posterior more than anterior) part of the default mode and frontal-parietal attentive-executive networks in relation to clinical presentation when observing very young children. This observation is in line with a possible altered developmental path of neural networks involving frontal regions [39] and with an age-related inversion of trends in functional connectivity in children with ASD, from initial overconnectivity to later under-connectivity [4], [40].

Although it is difficult to compare the results directly because of methodological differences (e.g., EEG vs. MEG, frequency bands, source estimation methods, and connectivity and graph-theory based approach), some evidence about brain connectivity derived by MEG in school age children converges in revealing that higher levels of ASD traits are related to brain under-connectivity and reduced efficiency [9], [12]. These result together with the present evidence of more severe ASD traits related to higher connectivity strength in pre-schoolers, support again the hypothesis that atypical ASD developmental patterns shift from intrinsic hyper-connectivity during the early childhood to hypo-connectivity during the pubertal period [4], [40].

Similar to other studies, we have found few significant correlations with ADOS severity scores [9], [12]. An explanation could be that many ADOS items are low-threshold, as they have been developed to identify core features of the disorder. Moreover, ADOS and ADI assess the autistic behaviours at different time scales. The ADOS-2 might not capture the behaviours that the child shows at home, due to the short time of observation, different environment, and an unfamiliar experimenter doing the assessment. The ADI asks about the traits as currently experienced and at previous years. On the other hand, parents are not usually trained to evaluate ASD manifestations accurately, especially in young children at first diagnostic assessment, and ADOS provides a direct view of the child by the clinician. The use of both a direct measure of the current child behaviour and the anamnestic interview is a point of strength of our work, as most studies evaluate clinical severity only with screening questionnaires filled by parents or with ADOS and screening parent reported questionnaires.

The present results should be interpreted in the light of some limitations. First, although similar to most functional connectivity investigations in children and adolescents, sample size is small. This is mainly due to the difficulty in organising effective pharmacological sedation procedures not interfering with the electroencephalographic signal, in particular with young children with neurodevelopmental disabilities. The administration of sedation was given at equal dosage/kg to all the patients to assure the control of this variable in influencing the acquired MEG signal and connectivity estimation. Moreover, we did not implemented comparison with a control group of typically developing children and/or with another developmental disability (e.g. children with global developmental delay without ASD), but intended to investigate the correlation between connectivity indices and the severity of autistic traits. The convenience sampling method did not allow sample balancing for sex, even if it is quite representative of male to female ratio in general population with ASD (4,3:1).

The present study has also important strengths with respect to previous investigations. In fact, we have studied very young children with wide range of global developmental level, still not exposed to medications, evidencing possible pathophysiological mechanism present at the onset of the disorder that could be generalized to children across the spectrum.

Moreover, the present study help strengthen the knowledge about the neurophysiological functional network of deeply studied young children with ASD in a narrow age range, extending the use of a valid and reliable functional connectivity method to this clinical population. Future studies implementing the same method on wider and stratified sample may further support functional connectivity as a possible biomarker of autism.

## Data Availability

All data produced in the present study are available upon reasonable request to the authors

## Acknowledgement

The study was supported by the Italian Ministry of Health (RRC) and by Pierfranco and Luisa Mariani foundation.

