## Supplementary Materials for "Correlation between autistic traits and brain functional connectivity in preschoolers with autism spectrum disorder: a resting state MEG study"

**Table S1 – Demographic and Neuropsychological data**

| **ID** | **sex** | **Age range months** | **GQ** | **ADI_Com** | **ADI_Intera** | **ADI_Intere** | **ADOS_Mod** | **ADOS_CSS** | **ADOS_SocAff** | **ADOS_RRB** |
| --- | --- | --- | --- | --- | --- | --- | --- | --- | --- | --- |
| 1 | M | 26-30 | 85 | 2 | 1,09 | 2 | 1 | 6 | 1,2 | 1,75 |
| 2 | M | 41-45 | 62 | 0,86 | 1,15 | 1,2 | 1 | 7 | 1 | 1,75 |
| 3 | M | 46-50 | 50 | 1,57 | 1,15 | 0,8 | 1 | 6 | 1,3 | 0,75 |
| 4 | M | 36-40 | 56 | 1,25 | 1,27 | 1,25 | 1 | 6 | 1,6 | 1,25 |
| 5 | M | 61-65 | 38 | 1,38 | 1,47 | 0,67 | 2 | 10 | 1,6 | 1,75 |
| 6 | F | 31-35 | 58 | 2 | 1,82 | 1,75 | 1 | 9 | 1,9 | 1,25 |
| 7 | M | 36-40 | 56 | 0,57 | 0,73 | 1 | 1 | 6 | 0,6 | 1,5 |
| 8 | M | 36-40 | 64 | 1,14 | 1,08 | 1 | 1 | 8 | 1,5 | 1 |
| 9 | M | 61-65 | 43 | 1,57 | 1,6 | 1 | 1 | 9 | 1,6 | 1,75 |
| 10 | M | 46-50 | 51 | 1,29 | 1,47 | 1,2 | 1 | 6 | 1,3 | 1,5 |
| 11 | M | 36-40 | 43 | 2 | 1,82 | 1,5 | 1 | 10 | 1,9 | 1,5 |
| 12 | M | 36-40 | 50 | 2 | 1,73 | 1 | 1 | 8 | 1,7 | 1,5 |
| 13 | M | 36-40 | 95 | 1 | 1 | 0,83 | 2 | 9 | 1,1 | 1,5 |
| 14 | F | 76-80 | 62 | 2 | 2 | 0,8 | 1 | 8 | 1,9 | 1,25 |
| 15 | M | 31-35 | 50 | 2 | 1,45 | 1 | 1 | 6 | 1,6 | 0,75 |
| 16 | F | 41-45 | 69 | 0,6 | 0,3 | 0,4 | 2 | 6 | 0,8 | 0,75 |

Legend: age months range: age in months, with a 6 months range. GQ: global quotient; ADI-Com: ADI Communication; ADI_Intere: ADI Restricted Interests; ADI_Intera: ADI Interaction; ADOS_Mod: ADOS Module (1: Non verbal or single words; 2: Phrases); ADOS_CSS: ADOS Calibrated Severity Score; ADOS_SocAff: ADOS Social Affect; ADOS_RRB: ADOS Restricted and Repetitive behaviors; M: male; F: Female.

**Table S2 Main clusters presenting increased FC-st at higher ADI Communication severity score**

| **ROI** | | **P-values** | **Percent. Cluster/ROI** | **LOCALIZATION** | **RSNetwork^1^** | **FUNCTION^2^** |
| --- | --- | --- | --- | --- | --- | --- |
|  | **β_Com1** | | | | | |
| Posterior Cingulate Gy_R | | 0,009 | 100% | Limbic_R | Default mode (posterior); Central Executive | Perception: somoesthesis, pain, gustation; Action: preparation, execution; Cognition: explicit memory, social cognition; Emotion processing. |
| Superior Occipital lobe_R | | 0,016 | 40% | O_R | Visual | Visual processing, motion perception |
| Cuneus_R | | 0,014 | 69,2% |  |  |  |
| Angular Gy_R | | 0,023 | 38,9% | PT_R | Default mode | Perception: motion vision; Cognition: language, working memory, attention, social cognition |
| Inferior Parietal Gy_R | | 0,028 | 81,8% | P_R | Default mode; Central Executive | Perception: motion, shape, somoesthesis, pain; Interoception; Cognition: explicit and working memory, reasoning, space, social cognition, attention |
| Superior Parietal Gy_R | | 0,033 | 94,4% | P_R | Central Executive | Action: immagination, preparation, execution, inhibition, learning; Cognition: reasoning, attention, working memory, space, soma, Perception: motion and shape vision |
| Postcentral Gy_R | | 0,039 | 51,5% | P_R | Sensorimotor | Perception: somesthesis, pain; Action: (speech) execution |
| Paracentral Lobule_R | | 0,042 | 87,5% | P_R | Sensorimotor | Interoception; Action: execution |
|  | **β_Com2** | | | | | |
| Superior Parietal Gy_L | | 0.034 | 87,5% | P_L | Central Executive | Action: immagination, preparation and execution; Cognition: attention, reasoning, working memory, somatic, space; Perception: motion and shape vision |
|  | **γ_Com** | | | | | |
| Middle cingulate Gy_R | | 0,015 | 40% | Limbic_Bil | Default mode (posterior); Central Executive | Perception: somoesthesis, pain, gustation; Action: preparation, execution; Cognition: explicit memory, social cognition; Emotion processing. |
| Middle cingulate Gy_L  Posterior cingulate Gy_R  Posterior cingulate Gy_L | | 0,003  0,008  0,001 | 50%  100%  80% |  |  |  |
| Inferior Occipital Lobe_L  Middle Occipital Lobe_L | | 0,000  0,004 | 50%  89,7% | O_L | Visual | Perception: vision, motion, shape; Cognition: space and language/orthography |
| Calcarine fissure and surranding ctx _R | | 0,003 | 41,7% | O_Bil | Visual | Perception: vision, motion, shape; Cognition: reasoning, shape, working memory |
| Superior Occipital lobe_R | | 0,008 | 63,6% |  |  |  |
| Cuneus_R | | 0,008 | 81,8% |  |  |  |
| Calcarine fissure and surranding ctx_L | | 0,003 | 60% |  |  |  |
| Superior Occipital Lobe_L | | 0,007 | 63,6% |  |  |  |
| Cuneus_L | | 0,007 | 81,8% |  |  |  |
| Lingual Gy_R | | 0,001 | 55,6% | OT_Bil | Visual | Perception: visual and shape processing |
| Lingual Gy_L | | 0,001 | 85,7% |  |  |  |
| Angular Gy_L | | 0,012 | 44,4% | PT_Bil | Default mode | Perception: motion vision; Cognition: language, working memory, attention, social cognition |
| Angular Gy_L | | 0,012 | 44,4% |  |  |  |
| Supramarginal Gy_R | | 0,016 | 50% | P_R | Salience | Interception; Perception: pain; Action: execution; Cognition: attention, social cognition |
| Precuneus_R | | 0,015 | 66,7% | P_Bil | Default mode; Parieto-Occipital | Perception: visual motion, Cognition: working memory, space, social cognition, explicit memory, language |
| Precuneus_L | | 0,018 | 78,6% |  |  |  |
| Inferior Parietal Gy_R | | 0,019 | 90,9% | P_Bil | Default mode; Central Executive | Perception: motion, shape, somoesthesis, pain; Interoception; Cognition: explicit and working memory, reasoning, space, social cognition, attention |
| Inferior Parietal Gy_L | | 0,017 | 55,6% |  |  |  |
| Superior Parietal Gy_R | | 0,026 | 100% | P_Bil | Central Executive | Action: immagination, preparation, execution, inhibition, learning; Cognition: reasoning, attention, working memory, space, soma, Perception: motion and shape vision |
| Superior Parietal Gy_L | | 0,024 | 100% |  |  |  |
| Postcentral Gy_R | | 0,033 | 66,7% | P_R | Sensorimotor | Perception: somesthesis, pain; Action: (speech) execution |
| Paracentral Lobule_R | | 0,039 | 100% | P_Bil | Sensorimotor | Interoception; Action: execution |
| Paracentral Lobule_L | | 0,040 | 77,8% |  |  |  |
| Precentral Gy_R | | 0,036 | 48,1% | M_R | Sensorimotor | Action: (speech) execution, motor learning; Perception: shape and motion vision |

Legend*: ROI, Region of Interest; Percent. Cluster/ROI: percentage of significant cluster’s nodes in a ROI; FC-st, Functional connectivity strength; RS, Resting-state; Gy, Gyrus; P, Parietal; T, Temporal; O, Occipital; L, Limbic; M, Motor; L, left; R, right; Bil, Bilateral.*

^1^Yeo et al., 2011; Doucet et al., 2019. ^2^ *Human Brainnetome Atlas* (Fan et al., 2016).

**Table S3 Main clusters presenting increased FC-st at higher ADI Interaction severity score**

| **ROI** | **P-values** | | **Percent. Cluster/ROI** | **LOCALIZATION** | **RSNetwork** | **FUNCTION** |
| --- | --- | --- | --- | --- | --- | --- |
|  | | **γ_Intera1** | | | | |
| Angular Gy_R | 0,009 | | 33,3% | PT_R | Default mode | Perception: motion vision; Cognition: language, working memory, attention, social cognition |
| Supramarginal Gy_R | 0,011 | | 50% | P_R | Salience | Interoception; Perception: pain; Action: execution; Congition: attention, social cognition |
| Inferior Parietal Gy_R | 0,014 | | 90,9% | P_R | Default mode; Central Executive | Perception: motion, shape, somoesthesis, pain; Interoception; Cognition: explicit and working memory, reasoning, space, social cognition, attention |
| Superior Parietal Gy_R | 0,029 | | 33,3% | P_R | Central Executive | Action: immagination, preparation, execution, inhibition, learning; Cognition: reasoning, attention, working memory, space, soma, Perception: motion and shape vision |
| Postcentral Gy_R | 0,030 | | 48,5% | P_R | Sensorimotor | Perception: somesthesis, pain; Action: (speech) execution |
| **γ_Intera2** | | | | | | |
| Superior Parietal Gy_L | 0,031 | | 62,5% | P_L | Central Executive | Action: immagination, preparation, execution; Cognition: reasoning, attention, working memory, space, soma; Perception: motion and shape vision |
| Paracentral lobule_R | 0,041 | | 50% | P_R | Sensorimotor | Interoception; Action: execution |
| **γ_Intera3** | | | | | | |
| Calcarine fissure and surrounding ctx_L | 0,030 | | 50% | O_L | Visual | Perception: vision, motion, shape |
| Lingual Gy_L | 0,017 | | 35,7% | OT_L | Visual | Perception: visual processing and memory |

Legend: *ROI, Region of Interest; Percent. Cluster/ROI: percentage of significant cluster’s nodes in a ROI; FC-st, Functional connectivity strenght; RS, Resting-state; Gy, Gyrus; P, Parietal; R, right.*

**Table S4 Main clusters presenting increased FC-st at higher ADI Interests severity score**

| **ROI** | **P-values** | | **Percent. Cluster/ROI** | **LOCALIZATION** | **RSNetwork** | **FUNCTION** |
| --- | --- | --- | --- | --- | --- | --- |
|  | | **β_Ints1** | | | | |
| Inferior Occipital Lobe_L | 0,003 | | 37,5% | O_L | Visual | Perception: shape vision; Cognition: orthography |
| Middle Temporal Gy_L | 0,019 | | 63,6% | T_L | Default mode; Central Executive | Perception: auditory; Cognition: explicit memory, language (semantic, phonology, speech, syntax), social cognition |
| Superior Temporal Gy_L | 0,033 | | 60% | T_L | Sensorimotor | Action: speech execution and motor learning; Perception: auditory; Cognition: language (phonology, semantic, speech), music; Emotion processing |
| Fusiform Gy_L | 0,003 | | 33,3% | T_L | Visual | Perception: visual perception: Cognition: language (semantich, speech, phonology, orthography), explicit memory; Emotion processing |
|  | | **β_Ints2** | | | | |
| Paracentral lobule_L | 0,041 | | 44,4% | P_L | Sensorimotor | Action: immagination and execution |
|  | | **γ_Ints** | | | | |
| Middle Temporal Gy_L | 0,018 | | 34,1% | T_L | Default mode; Central Executive | Perception: auditory; Cognition: explicit memory, language (semantic, phonology, speech, syntax), social cognition |
| Superior Temporal Gy_L | 0,021 | | 45% | T_L | Sensorimotor | Action: speech execution and motor learning; Perception: auditory; Cognition: language (phonology, semantic, speech), music; Emotion processing |

Legend: *ROI, Region of Interest; Percent. Cluster/ROI: percentage of significant cluster’s nodes in a ROI; FC-st, Functional connectivity strenght; RS, Resting-state; Gy, Gyrus; P, Parietal; T, Temporal; O, Occipital; L, left.*

**Table S5 Main clusters presenting increased FC-st at higher ADOS Social Affect severity score**

| **ROI** | **P-values** | | **Percent. Cluster/ROI** | **LOCALIZATION** | **RSNetwork** | **FUNCTION** |
| --- | --- | --- | --- | --- | --- | --- |
|  | | **γ_Soc1** | | | | |
| Inferior Parietal Gy_R | 0,025 | | 63,6% | P_R | Default mode; Central Executive | Perception: motion, shape, somoesthesis, pain, interoception; Cognition: explicit and working memory, reasoning, space, social cognition, attention |
| Supramarginal Gy_R | 0,021 | | 30% | P_R | Salience | Interoception; Perception: pain; Action: execution; Congition: attention, social cognition |
|  | | **γ_Soc2** | | | | |
| Lingual Gy_L | 0,020 | | 35,7% | OT_L | Visual | Perception: visual processing |
| Calcarine fissure and surroundin ctx_L | 0,020 | | 35% | O_L | Visual | Perception: motion and shape vision |

Legend: *ROI, Region of Interest; Percent. Cluster/ROI: percentage of significant cluster’s nodes in a ROI; FC-st, Functional connectivity strenght; RS, Resting-state; Gy, Gyrus; P, Parietal; O, Occipital; L, left; R, right.*

**Table S6. Seed-based analyses and association to ADI Communication severity score**

| **AREA** | **LOCALIZATION** | **PERC. CLUSTERS/ROIS** |
| --- | --- | --- |
| **β_Com1** | | |
| **Seed 1** |  |  |
| Paracentral lobule R-L | P_Bil | 75%-89% |
| Postcentral gyrusR | P_R | 33% |
| Superior Parietal gyrusL | P_L | 63% |
| Angular gyrusL | PT_L | 67% |
| Supplementary Motor area R-L | F_Bil | 100%-100% |
| Middle Frontal gyrus, OrbitalR | F_R | 71% |
| Superior Frontal gyrus medial and orbital R | F_R | 33%; 33% |
| Inferior Occipital lobe R-L | O_Bil | 100% - 50% |
| Fusiform gyrusR | O_R | 84% |
| Superior Occipital lobeR | O_R | 50.0% |
| Calcarine fissure and surrounding cortexR | O_R | 83% |
| CuneusL | O_L | 36% |
| Lingual gyrusL | OT_L | 100% |
| Temporal pole, uperior-middle-inferior Temporal gyrusR | T_R | 63% - 70% - 65% |
| Cingulate gyrus, Posterior part R-L | Limbic_Bil | 50% - 40% |
| ParahippocampusR | Limbic_R | 70.0% |
| AmygdalaR | Limbic_R | 100% |
| **Seed 2** |  |  |
| Gyrus RectusL | F_L | 75% |
| **β_Com2** | | |
| **Seed 1** |  |  |
| Precentral gyrus R-L | P_Bil | 93% - 31% |
| Postcentral gyrus R-L | P_Bil | 76% - 32% |
| Paracentral lobuleR | P_R | 50% |
| Inferior Parietal gyrusR | P_R | 82% |
| Angular gyrusR | PT_L | 61% |
| Supramarginal gyrusR | F_R | 60% |
| Supplementary Motor area R-L | F_Bil | 100% - 88% |
| Superior Frontal gyrus R-L | F_Bil | 81% - 37% |
| Superior Frontal gyrus, Orbital R-L, Medial R-L | F_Bil | 100%-33%-73%-79% |
| Superior Frontal gyrus, Medial OrbitalL | F_L | 33% |
| Middle Frontal gyrus, OrbitalR | F_R | 100% |
| Middle Frontal gyrusR | F_L | 85% |
| Inferior Frontal gyrus, Triangular, Opercular, Orbital R | F_R | 74%-85%-92% |
| Gyrus Rectus R-L | F_Bil | 100%-50% |
| Superior-Middle-Inferior Temporal gyrus R | T_R | 89%-86%-50% |
| Rolandic operculumR | FPT_R | 91% |
| Temporal pole, Superior-Middle temporal gyrus R | T_R | 88%-100% |
| Fusiform gyrusR | O_R | 59% |
| Cingulate gyrus, Anterior part R-L | Limbic_Bil | 40%-89% |
| Cingulate gyrus, Middle part R-L | Limbic_Bil | 80%-63% |
| ParahippocampusR | Limbic_R | 100% |
| HippocampusR | Limbic_R | 100% |
| InsulaR | Limbic_R | 93% |
| AmygdalaR | Limbic_R | 100% |
| **Seed 2** |  |  |
| Lingual gyrusL | O_L | 100% |
| Calcarine fissure and surrounding cortexL | O_L | 55% |
| Inferior Occipital lobeL | O_L | 88% |
| Fusiform gyrusL | O_L | 47% |
| Cingulate gyrus, Posterior partL | O_L | 40% |
| **Seed 3** |  |  |
| Inferior Frontal gyrus, OpercularL | F_L | 43% |
| Rolandic operculumL | FTP_L | 60% |
| **γ_Com** | | |
| **Seed 1** |  |  |
| Postcentral gyrus R-L | P_Bil | 89%-82% |
| Paracentral lobule R-L | P_Bil | 100%-100% |
| Superior R-L, Inferior R-L Parietal gyrus | P_Bil | 50%-50%-55%-61% |
| Supramarginal gyrus R-L | P_Bil | 30%-50% |
| PrecuneusL | P_L | 46% |
| Angular gyrus R-L | PT_ Bil | 61%-89% |
| Precentral gyrus R-L | Motor_Bil | 100%-84% |
| Supplementary Motor area R-L | F_Bil | 100%-100% |
| Middle Frontal gyrus R-L , and orbital R | F_Bil | 68%-44%;71% |
| Superior Frontal gyrus, medial, orbital, medial orbitalR | F_R | 65%; 53%; 33%; 40% |
| Superior Frontal gyrus, medial, orbital, medial orbitalL | F_R | 52%; 76%; 67%; 83% |
| Gyrus rectus R-L | F_Bil | 100%-63% |
| Inferior Frontal gyrus, Triangular, opercular, orbitalR | F_R | 53%; 54%; 50% |
| Rolandic operculum R-L | FPT_Bil | 82%-60% |
| Middle temporal gyrus R-L and temporal pole R | T_Bil | 49%; 80% |
| Superior Temporal gyrus and temporal poleR | T_R | 70%; 50% |
| Inferior Temporal gyrusR | T_L | 65% |
| Lingual gyrus R-L | OT_Bil | 89%-100% |
| Fusiform gyrus R-L | O_Bil | 95%-53% |
| Inferior Occipital lobe R-L | O_Bil | 100%-100% |
| Middle Occipital lobe R-L | O_Bil | 71%-97% |
| Superior Occipital lobe R-L | O_Bil | 70%-73% |
| Calcarine fissure and surrounding cortex R-L | O_Bil | 100%-90% |
| Cuneus R-L | O_Bil | 31%-36% |
| Cingulate gyrus, posterior, middle and anterior partR | Limbic_R | 75%; 87%; 40% |
| Cingulate gyrus, posterior, middle and anterior partL | Limbic_L | 60%; 88%; 84% |
| InsulaR | Limbic_R | 71% |
| HippocampusR | Limbic_R | 100% |
| ParahippocampusR | Limbic_R | 90.0% |
| AmygdalaR | Limbic_R | 100% |

  Legend: ROIs, Regions of Interest; Gy, Gyrus; P, Parietal; T, Temporal; O, Occipital; F, Frontal; M, Motor; L, left; R, right; SMA: Supplementary Motor Area.

**Table S7 Seed- based analysis and association to ADI Interaction severity score**

| **AREA** | **LOCALIZATION** | **PERC. CLUSTERS/ROIS** |
| --- | --- | --- |
| **γ_Intera1** | | |
| **Seed 1** |  |  |
| Paracentral lobule R-L | P_Bil | 38%-100% |
| Postcentral gyrusL | P_L | 71% |
| Superior, Inferior Parietal gyrus L | P_L | 31%-83% |
| Angular gyrusL | PT_L | 100% |
| Precentral gyrusL | Motor_L | 63% |
| Supplementary Motor area R-L | F_Bil | 43%-50% |
| Superior Frontal gyrus and Medial R-L | F_Bil | 74%-37% ; 33%-32% |
| Middle Frontal gyrusR | F_R | 80.0% |
| Inferior Frontal gyrus, OpercularR | F_R | 54% |
| Lingual gyrusL | OT_L | 93% |
| Superior, Middle, Inferior Occipital lobe L | O_L | 45%-69%-36% |
| Calcarine fissure and surrounding cortexL | O_L | 45% |
| InsulaR | Limbic_R | 64% |
| Cingulate gyrus, Middle partL | Limbic_L | 63% |
| **Seed 2** |  |  |
| Temporal pole, Superior-Middle-Inferior temporal gyrusR | T_R | 38%-50%-35% |
| Fusiform gyrusR | O_R | 68% |
| HippocampusR | Limbic_R | 57% |
| ParahippocampusR | Limbic_R | 80.0% |
| AmygdalaR | Limbic_R | 100% |
| **γ_Intera2** | | |
| **Seed 1** |  |  |
| Right Precentral gyrus | P_R | 44% |
| Paracentral lobule R-L | P_Bil | 75%-44% |
| Postcentral gyrusR | P_R | 46% |
| Inferior Parietal gyrusR | P_R | 64% |
| Angular gyrusR | PT_R | 67% |
| Supramarginal gyrusR | P_R | 80% |
| Superior Frontal gyrusR | F_R | 32% |
| Superior Frontal gyrus, Medial R-L | F_Bil | 40%-32% |
| Middle Frontal gyrusR | F_R | 50% |
| Rolandic operculumR | FTP_R | 82% |
| Superior Temporal gyrusR | T_R | 74% |
| Temporal pole, Superior Temporal gyrusR | T_R | 50% |
| Middle temporal gyrusR | T_R | 41% |
| Temporal pole, Middle temporal gyrusR | T_R | 50% |
| Inferior Temporal gyrusR | T_R | 38% |
| Fusiform gyrusR | O_R | 47% |
| Cingulate gyrus, Anterior R-L, Middle R-L part | Limbic_Bil | 40%-68%; 80%-31% |
| ParahippocampusR | Limbic_R | 60% |
| HippocampusR | Limbic_R | 43% |
| InsulaR | Limbic_R | 36% |
| AmygdalaR | Limbic_R | 100% |
| **Seed 2** |  |  |
| Lingual gyrusL | O_L | 86% |
| Middle Occipital lobeL | O_L | 41% |
| Calcarine fissure and surrounding cortexL | O_L | 55% |
| CuneusL | O_L | 54% |
| Superior Occipital lobeL | O_L | 45% |
| **γIntera3** | | |
| **Seed 1** |  |  |
| Postcentral gyrusR | P_R | 61% |
| Postcentral gyrusL | P_L | 47% |
| Paracentral lobuleR | P_R | 100% |
| Paracentral lobuleL | P_L | 89% |
| Precentral gyrusR | Motor_R | 56% |
| Superior Parietal gyrusL | P_L | 75% |
| Inferior Parietal gyrusR | P_R | 73% |
| Inferior Parietal gyrusL | P_L | 44% |
| Supramarginal gyrusR | P_R | 60% |
| Angular gyrusR | PT_R | 61% |
| Middle Frontal gyrsusR | F_R | 38% |
| Superior Frontal gyrusR | F_R | 35% |
| Lingual gyrusL | OT_L | 79% |
| Calcarine fissure and surrounding cortexL | O_L | 55% |
| Superior Occipital lobeL | O_L | 36% |
| Cingulate gyrus, Posterior partR | Limbic_R | 75% |
| Cingulate gyrus, Middle partR | Limbic_R | 67% |
| Cingulate gyrus, Middle partL | Limbic_L | 69% |
| **Seed 2** |  |  |
| Right Temporal pole, Middle temporal gyrus | T_R | 50% |
| Right Temporal pole, Superior Temporal gyrus | T_R | 50% |
| Right Parahippocampus | Limbic_R | 30% |
| **Seed 3** |  |  |
| Fusiform gyrus | O_R | 37% |

Legend: ROIs, Regions of Interest; Gy, Gyrus; P, Parietal; T, Temporal; O, Occipital; F, Frontal; M, Motor; L, left; R, right; SMA: Supplementary Motor Area.

**Table S8 Seed-based analysis and association to ADI Interests severity score**

| **AREA** | **LOCALIZATION** | **PERC. CLUSTERS/ROIS** |
| --- | --- | --- |
| **β_Ints1** | | |
| **Seed 1** |  |  |
| Paracentral lobule R-L | P_Bil | 100% |
| Postcentral gyrus L | P_L | 53% |
| Supplementary Motor area R-L | F_Bil | 50%-54% |
| Precentral gyrus L | Motor_L | 50.0% |
| Cingulate gyrus, Middle part R-L | Limbic_Bil | 40.0% |
| **Seed 2** |  |  |
| Superior Frontal gyrus, Medial Orbital and orbitalR | F_R | 40%; 67% |
| Inferior Frontal gyrus orbital and gyrus rectusL | F_L | 33%; 62% |
| Superior Frontal gyrus orbitalL | F_L | 67% |
| Temporal pole middle temporal gyrusL | T_L | 57% |
| Temporal pole superior Temporal gyrusL | T_L | 50.0% |
| Fusiform gyrusL | O_L | 33% |
| ParahippocampusL | Limbic_L | 50.0% |
| AmygdalaL | Limbic_L | 100% |
| **Seed 3** |  |  |
| Middle Frontal gyrus and orbitalL | F_L | 35%, 43% |
| **Seed 4** |  |  |
| Superior Temporal gyrusL | T_L | 55% |
| **Seed 5** |  |  |
| Inferior Frontal gyrus, Orbital | F_R | 58% |
| **β_Ints2** | | |
| **Seed 1** |  |  |
| Superior Frontal gyrus, OrbitalL | F_L | 100% |
| Middle Frontal gyrus and OrbitalisL | F_L | 53%; 86% |
| Inferior Frontal gyrus Triangular, Orbital and opercularL | F_L | 62%; 83%; 86% |
| Rolandic operculumL | FTP_L | 40.0% |
| Superior Temporal gyrus and temporal poleL | T_L | 65%; 90% |
| Middle temporal gyrus and temporal poleL | T_L | 75%, 100% |
| Lingual gyrusL | OT_L | 36% |
| Fusiform gyrusL | O_L | 93% |
| Middle Occipital lobeL | O_L | 45% |
| Inferior Occipital lobeL | O_L | 75% |
| HippocampusL | Limbic_L | 80.0% |
| ParahippocampusL | Limbic_L | 75% |
| AmygdalaL | Limbic_L | 100% |
| **γ_Intera1** | | |
| **Seed 1** |  |  |
| Paracentral lobuleR | P_R | 75% |
| Paracentral lobuleL | P_L | 89% |
| Supplementary Motor areaR | F_R | 36% |
| Supplementary Motor areaL | F_L | 38% |
| Precentral gyrusL | Motor_L | 50.0% |
| Postcentral gyrusL | P_L | 35% |
| **Seed 2** |  |  |
| Fusiform gyrusR | O_R | 68% |
| Fusiform gyrusL | O_L | 33% |
| Inferior Frontal gyrus orbital and gyrus rectusL | F_L | 42%; 50% |
| Middle Frontal gyrus orbitalL | F_L | 71% |
| Superior Frontal gyrus medial-orbital and orbitalL | F_L | 83%; 100% |
| Temporal pole middle temporal gyrusL | T_L | 57% |
| Temporal anterior (pole) superiorL | T_L | 50.0% |
| Cingulate gyrus anterior partL | Limbic_L | 37% |
| HippocampusL | Limbic_L | 40.0% |
| ParahippocampusL | Limbic_L | 50.0% |
| AmygdalaL | Limbic_L | 100% |
| **Seed 3** |  |  |
| Superior Temporal gyrusL | T_L | 45% |

  Legend: ROIs, Regions of Interest; Gy, Gyrus; P, Parietal; T, Temporal; O, Occipital; F, Frontal; M, Motor; L, left; R, right; SMA: Supplementary Motor Area.

**Table S9 Seed-based analysis and association to ADOS Social Affect severity score**

| **AREA** | **LOCALIZATION** | **PERC. CLUSTERS/ROIS** |
| --- | --- | --- |
| **γ_Soc1** | | |
| **Seed 1** |  |  |
| Lingual gyrus R-L | OT_Bil | 39%-57% |
| Fusiform gyrusR | O_R | 47% |
| Inferior Occipital lobeR | O_R | 40.0% |
| Middle Occipital lobeL | O_L | 45% |
| Calcarine fissure and surroun cortexL | O_L | 40.0% |
| Inferior Temporal gyrusR | T_R | 54% |
| **Seed 2** |  |  |
| Middle Frontal gyrusR | F_R | 38% |
| **γ_Soc2** | | |
| **Seed 1** |  |  |
| Postcentral gyrus R-L | P_Bil | 42%-35% |
| Precuneus R-L | P_Bil | 57%-32% |
| Paracentral lobule R-L | P_Bil | 88%-78% |
| Superior Parietal gyrus R-L | P_Bil | 50%-94% |
| Inferior Parietal gyrus R-L | P_Bil | 82%-33% |
| Supramarginal gyrusR | P_R | 40.0% |
| Calcarine fissure and surrounding cortexL | O_L | 75% |
| Superior Occipital lobeL | O_L | 55% |
| Middle Occipital lobeL | O_L | 52% |
| Lingual gyrus R-L | OT_Bil | 50%-57% |
| Angular gyrusR | O_R | 56% |
| Cingulate gyrus, Middle part R-L | Limbic_Bil | 33%-56% |
| Cingulate gyrus, Posterior partR | Limbic_R | 100% |
| **Seed 1** |  |  |
| Temporal pole, Middle temporal gyrusR | T_R | 50.0% |
| Temporal pole, Superior Temporal gyrusR | T_R | 38% |
| Fusiform gyrusR | O_R | 42% |
| ParahippocampusR | Limbic_R | 40.0% |

Legend: ROIs, Regions of Interest; Gy, Gyrus; P, Parietal; T, Temporal; O, Occipital; L, left; R, right.

**Supplementary results**

**Seed Analysis**

Increased cluster FC-st indicates that the oscillatory activity (within a given frequency band) of the cluster regions is more synchronously paired with activity from all across the brain. However, in order to specifically identifying which connection drove such an effect, we performed a seed-based analysis. In this seed-based correlation analysis, we identified the specific connections (seed clusters) of each of the main clusters with the rest of the brain that were significantly modulated by severity scores at ADI-R and ADOS-2 scales

*ADI Communication*

One significant seed (s) cluster emerged for the main cluster in gamma band (sγCom), spread out in several bilateral fronto-parietal, temporo-occipital and mainly right-lateralized limbic ROIs. As for beta band, two significant seed clusters emerged for βCom1 (s1βCom1 and s2βCom1) and three seed clusters emerged for βCom2 (s1βCom2, s2βCom2 and s3βCom2). s1βCom1 seed cluster was highly diffused in several bilateral regions the brain encompassing frontal, parietal, temporal, occipital and limbic ROIs, while s2βCom1 was more localized in the left frontal gyrus rectus. s1βCom2 seed cluster is very diffused in the brain, reaching several bilateral parietal, temporal, frontal and limbic ROIs, while s2βCom2 and s3βCom2 seed clusters are left lateralized respectively in the occipital lobe and in the inferior frontal gyrus and the rolandic operculum. The detailed list of areas belonging to the seed clusters is shown in Fig 1, right panels and Supplementary Table S6.

*ADI Interaction*

Two significant seed clusters for main clusters in gamma band γIntera1 and γIntera2 emerged (s1γIntera1 and s2γIntera1 for γIntera1, s1γIntera2 and s2γIntera2 for γIntera2), while three significant seed clusters emerged for γIntera3 (s1γIntera3, s2γIntera3 and s3γIntera3). S1γIntera1 involves regions mainly belonging to the left frontoparietal and occipital ROIs, while s2γIntera1 is mainly localized in right temporal and limbic regions. s1γIntera2 is diffused in bilateral parietal, frontal, temporal and limbic ROIs, while s2γIntera2 is left localized in occipital ROIs. S1γIntera3 is diffused mainly in parietal and some frontal, occipital and limbic ROIs, while s2γIntera3 and s3γIntera3 are right lateralized respectively in mesial temporal and limbic regions and fusiform gyrus. The detailed list of areas belonging to these clusters is shown in in Fig 2, right panels and Supplementary Table S7.

*ADI Restricted Interests*

Three significant seed clusters have been identified for the main cluster in gamma band (s1γInts; s2γInts and s3γInts for γInts); s1γInts is located in fronto-parietal sensorimotor areas around the central sulcus, s2γInts is left lateralized and located in more anterior frontal areas and in inferior and middle temporal and limbic ROIs, s3γInts is localized in the left superior temporal gyrus. Moreover, five and one significant secondary clusters for the first and the second primary cluster in beta band emerged, respectively (s1βints1, s2βints1, s3βints1, s4βints1 and s5βints1for βints1, and s2βints2 for βints2). s1βints1 is located in sensorimotor fronto-parietal regions around the central sulcus and extends to the middle cingulate cortex, s2βints1 is left lateralized and involves the fusiform gyrus, anterior frontal areas, middle temporal and limbic ROIs, s3βints1, s4βints1 and s5βints1 are smaller second clusters located in the left superior frontal gyrus, left superior temporal gyrus and right inferior frontal gyrus, respectively. s2βints2 is left lateralized and frontal, occipito-temporal and limbic ROIs. The detailed list of areas belonging to these clusters is shown in Fig 3, right panels and Supplementary Table S8.

*ADOS Social Affect*

Two significant seed clusters emerged for each main cluster in the gamma band (s1γSoc1 and s2γSoc1 for γSoc1, s1γSoc2 and s2γSoc2 for γSoc2). s1γSoc1 and s2γSoc1 are located posteriorly in bilateral occipital lobe and anteriorly in the right middle frontal gyrus, respectively. s1γSoc2 is located in the bilateral parietal (from the paracentral lobule to the inferior parietal and supramarginal gyri), occipital and cingulate cortices and s2γSoc2 is located in the right temporal lobe, fusiform gyrus and parahippocampus. The detailed list of areas belonging to these clusters is shown in Fig 4.right panels and Supplementary Table S9.
